# Connectometry reveals differing associations of cortisol and PACAP with dorsal cingulum microstructure in posttraumatic stress

**DOI:** 10.1101/2025.06.05.25329085

**Authors:** Steven J. Granger, Sydney A. Jobson, Caitlin Ravichandran, Quentin Devignes, Eylül Akman, Jennifer U. Blackford, Victor May, Sayamwong E. Hammack, William A. Carlezon, Kerry J. Ressler, Scott L. Rauch, Isabelle M. Rosso

## Abstract

Posttraumatic stress disorder (PTSD) has been associated with altered arousal regulation and dysfunction of the hypothalamic-pituitary-adrenal axis, including changes in circulating cortisol and pituitary adenylate cyclase-activating polypeptide (PACAP). Both stress-related hormones affect extended amygdala to medial prefrontal cortex (mPFC) circuit functioning, but it is unclear whether they relate to white matter microstructure connecting these regions. We examined this question in 139 trauma-exposed adults (81 female; ages 19-54) who completed the Clinician-Administered PTSD Scale, a blood draw, and diffusion magnetic resonance imaging. White matter integrity was assessed in tracts connecting the extended amygdala to mPFC, including the uncinate fasciculus, frontal parahippocampal cingulum, and bed nucleus of the stria terminalis to mPFC projections. We used both tract-average fractional anisotropy (FA) to assess the global integrity of these white matter tracts and restricted connectometry to identify spatially localized associations along specific tract segments. Neither cortisol nor PACAP levels were associated with tract-average FA in any tract. However, connectometry, using a stringent statistical T-threshold revealed distinct, region-specific associations within the dorsal cingulum: higher cortisol levels were associated with lower FA (*FDR*=.002), whereas higher PACAP levels were associated with higher FA (*FDR*=.01). These localized FA alterations were not significantly associated with symptom severity. These findings suggest that cortisol and PACAP levels have differing associations with microstructural integrity of the dorsal cingulum, a region implicated in emotional regulation. These results highlight how distinct stress hormone pathways may differentially impact white matter organization in PTSD and demonstrate the utility of connectometry for detecting regionally specific brain-biomarker relationships.

## INTRODUCTION

Posttraumatic stress disorder (PTSD) is characterized by distressing and persistent symptoms, including altered arousal and reactivity following trauma exposure [1]. Dysregulation of the hypothalamic-pituitary-adrenal (HPA) axis, including changes in cortisol and pituitary adenylate cyclase-activating polypeptide (PACAP) levels, has been associated with PTSD diagnosis [2–5] and symptom severity [5–8]. These peripherally detected hormones are implicated in stress reactivity and PTSD symptom severity, particularly hyperarousal symptoms [5, 9]. Recent preclinical findings further suggest that these stress molecules exert differential effects on arousal processes, including sleep architecture [10]. Functionally, PACAP and cortisol influence brain circuits central to the stress response and PTSD, including those connecting the extended amygdala to the medial prefrontal cortex (mPFC) [11–13]. Major white matter tracts support these pathways, including the uncinate fasciculus (UF), the frontal parahippocampal cingulum, and fibers connecting the bed nucleus of the stria terminalis (BNST) with the mPFC. However, to our knowledge, no prior study has examined associations of both cortisol and PACAP with white matter integrity of these circuits, including in PTSD.

White matter integrity within amygdala-mPFC tracts has been variably associated with PTSD symptoms [14–17] and cortisol levels [18, 19]. Lower UF integrity has been associated with PTSD and hyperarousal symptoms in both diagnosed and subthreshold samples [14, 16]. Similarly, lower UF integrity has been linked to higher cortisol levels in adolescents [12] and in adults with stress-related pathologies, including major depressive disorder [18]. Associations between cingulum integrity and PTSD symptom severity or cortisol are less consistent, with some studies reporting positive [19] and others negative [15] associations, potentially related to comorbid conditions [20].

The BNST, an extended amygdala structure, plays a key role in sustained fear and hyperarousal [21], core features of PTSD. Corticotropin-releasing factor receptor 1 and PACAP receptor 1 (PAC1), both implicated in PTSD [5, 22], are expressed in the BNST and the mPFC, suggesting this pathway may be influenced by stress hormone signaling [23–25]. While altered BNST–mPFC functional connectivity have been reported during threat processing in individuals with PTSD [26], neither the structural integrity of this white matter circuit nor its relationship with stress hormone signaling has been examined. Investigating associations of cortisol and PACAP levels with white matter microstructure of this pathway may uncover novel biomarkers of PTSD-related arousal phenotypes.

In summary, cortisol and PACAP are implicated in PTSD pathophysiology. Altered white matter integrity of tracts connecting the amygdala to the mPFC, particularly the UF and frontal parahippocampal cingulum, have been linked to PTSD symptoms and cortisol levels. However, the direction of associations with cortisol has been inconsistent, and the relationship between PACAP and the integrity of these white matter pathways has not been examined in PTSD. Additionally, integrity of BNST–mPFC white matter has yet to be investigated in relation to PTSD or these stress hormones. To address these gaps, we investigated whether circulating cortisol and PACAP levels were associated with tract-average fractional anisotropy (FA), a marker of white matter integrity [27], in the UF, frontal parahippocampal cingulum, and BNST–mPFC tracts in trauma-exposed adults with PTSD symptoms. Because tract-average FA cannot detect associations localized to specific segments of white matter tracts, we also conducted connectometry [28], a data-driven method that can identify localized white matter segments where FA correlates with hormone levels. Finally, we evaluated the clinical significance of these findings by testing whether FA in hormone-associated segments predicted PTSD symptom severity (total and hyperarousal), PTSD diagnosis, and depression symptom severity.

## MATERIALS AND METHODS

### Participants and Procedure

One hundred and eighty-one (181) trauma-exposed adults were recruited from the greater Boston area to participate in a study examining stress pathways in PTSD [11, 29]. As previously described [11, 29], participants completed a fasting blood draw and a magnetic resonance imaging (MRI) session. Inclusion required DSM-5 Criterion A trauma exposure and meeting criteria for at least two of the four PTSD symptom clusters (B-E), assessed by the Clinician-Administered PTSD Scale for DSM-5 (CAPS-5) [30]. Table S1 shows trauma exposure types from the Life Events Checklist for DSM-5 [31]. Exclusion criteria are detailed in the Supplement. Forty-two participants were excluded due to missing or poor-quality diffusion imaging data or unavailable blood samples, yielding a final analytic sample of 139 (Table 1). All procedures were approved by the Mass General Brigham Human Research Committee, and participants provided written informed consent.

**Table 1.** Participant demographic and clinical characteristics (N=139)

| Characteristic | N (%) or Mean (SD) |
| --- | --- |
| Age (years) | 30.18 (8.58) |
| **Sex** |  |
| Female | 81 (58%) |
| Male | 58 (42%) |
| **Race** |  |
| Asian | 12 (8%) |
| Black | 14 (10%) |
| Native American or Alaskan Native | 1 (1%) |
| White | 93 (67%) |
| Bi-/multiracial | 10 (7%) |
| Unknown or not reported | 9 (7%) |
| **Ethnicity** |  |
| Hispanic | 24 (17%) |
| Non-Hispanic | 115 (83%) |
| **Gender** |  |
| Female | 78 (56%) |
| Male | 54 (39%) |
| Non-binary or gender non-conforming | 7 (5%) |
| **PACAP (pM)** | 903.03 (858.91) |
| **Cortisol (µg/dL)** | 11.05 (4.97) |
| **PTSD Diagnosis** |  |
| Yes | 89 (64%) |
| No | 50 (36%) |
| **CAPS-5 Total Score** | 28.33 (10.44) |
| **BDI-II Total Score** | 20.74 (10.18) |

*Note:* PACAP: pituitary adenylate cyclase-activating polypeptide; PTSD: posttraumatic stress disorder; CAPS-5: Clinician-Administered PTSD Scale for DSM-5; BDI-II: Beck Depression Inventory, Version II; SD: standard deviation

### Clinical Interviews and Questionnaires

The CAPS-5 [30], administered by doctoral-level psychologists, was used to assess DSM-5 PTSD diagnosis, total symptom severity, and hyperarousal symptom severity (Cluster E). Depression symptom severity was assessed using the 21-item Beck Depression Inventory-II (BDI-II) [32]. For both measures, higher scores indicate greater symptom severity.

### Cortisol and PACAP

Fasting blood draws were scheduled between 8:00 AM and 10:00 AM. Following collection, blood in EDTA tubes was centrifuged at 3500 rpm for 15 minutes for plasma separation. Serum samples were allowed to clot for at least 30 minutes before being centrifuged under the same conditions. Aliquots were extracted and then stored at −80°C until shipment for analysis. Cortisol was assayed from serum at the Center for Research in Reproduction Ligand Assay and Analysis Core at the University of Virginia School of Medicine. Serum samples were processed (Kit #L2KCO2, Cat #10381476, Siemens Healthcare Diagnostics, Los Angeles, CA) using an IMMULITE 2000 Immunoassay System (Siemens Healthcare Diagnostics, Los Angeles, CA) in three batches. Plasma samples were prepared for PACAP-38 immunoassay as described previously [5, 11, 29, 33]. All PACAP-38 measurements were performed at the University of Vermont, Larner College of Medicine, using double antibody sandwich ELISA immunoassays (Cat #HUFI02692, AssayGenie, Dublin, Ireland) in two batches. Ten samples had undetectable PACAP levels, resulting in a subsample of 129 participants for PACAP analyses.

### MRI Data Acquisition

Neuroimaging was conducted at the McLean Imaging Center on a 3.0 Tesla Siemens Prisma Scanner with a 64-channel head coil, using either the Human Connectome Project (HCP) Adult Lifespan (*n* = 126) or the HCP Young Adult (*n* = 13) protocol, which were developed to be compatible [34]. Each included T1-weighted MPRAGE and high-angular, multi-shell whole-brain diffusion-weighted imaging sequences. Full imaging acquisition parameters are provided in the Supplement. Identical diffusion sequences with opposite-phase encoding enabled correction for susceptibility-induced distortions.

### Diffusion MRI Processing

Opposite phase-encoding B0s were merged and processed with FSL’s *topup* to estimate susceptibility-induced off-resonance fields [35, 36]. These outputs were fed into FSL’s *eddy* tool to correct for motion, eddy current artifacts, and magnetization-induced susceptibility distortions [37]. Diffusion data were reconstructed with Q-Space Diffeomorphic Reconstruction (QSDR) in DSI Studio. Scans were filtered for template fit (see Supplement for image processing details), yielding 139 usable datasets. QSDR [41] was conducted to generate the spin distribution function with a diffusion sampling length ratio of 1.3, enabling visualization of crossing fibers at the corpus callosum and corticospinal tract intersection. QSDR was conducted with an output resolution of 1.25 mm^3^. Accuracy of b-table orientation was verified by comparing fiber orientations against a population-averaged template [42]. All usable diffusion MRI scans were included in the connectometry database.

### Tractography Procedures

Tractography was performed using a diffusion template derived from 1065 young adults [38]. This group-level approach derives tract-average FA values and has been used to examine associations between stress reactivity and cingulum and UF integrity [39], as well as to distinguish adjacent white matter tracts (e.g., terminalis and fornix) [40]. Given these precedents, we employed this method to delineate the BNST–mPFC pathway, which lies near and partially overlaps with the anterior thalamic radiation (Supplement). FA values were calculated from the connectometry database using *b-*value lower than 1750 s/mm² and used for subsequent analyses.

#### Automated Tractography for Major White Matter Bundles

The left and right UF and frontal parahippocampal cingulum tracts were delineated using an automated, augmented tractography function [43] that combines deterministic fiber tracking [44] with a tract-to-region atlas [38]. For this study, tracts are defined as large white matter bundles composed of groups of individual streamlines. Notably, this quantitative anisotropy-informed approach has been shown to eliminate false positive streamlines more effectively than 96 alternative methods [45]. Additional details regarding automated tractography are in the Supplement. Because BNST–mPFC white matter tracts are less well characterized, we developed a custom tractography protocol to delineate these tracts (see Supplement; Figure S1–3). Figure S4 shows the resulting reconstructions for the bilateral UF, frontal parahippocampal cingulum, and BNST–mPFC tracts. Tract-average FA values were extracted for each tract from the connectometry database.

#### Restricted Connectometry

Connectometry [28] and correlational tractography [46] were used to identify white matter segments within the UF, frontal parahippocampal cingulum, and BNST–mPFC pathway that exhibit microstructural associations with cortisol or PACAP levels. *Connectometry* refers to the statistical framework that calculates T-statistics for associations between a study variable and diffusion metrics, while *correlational tractography* refers to the tractography method used to map white matter segments with statistically significant associations. These methods reveal segments of white matter affected by chronic stroke lesions, making it a validated method to identify areas of white matter with structural abnormality [47].We refer to our data-driven analytic approach as *restricted connectometry*, as analyses were constrained to a binary mask (or region of interest [ROI]) encompassing the bilateral UF, frontal parahippocampal cingulum, and BNST–mPFC tracts (Figure S5). Spearman correlations were computed between blood hormone levels and adjusted FA values, controlling for age, sex, motion, draw time, and assay batch. Correlational tractography was performed using the resulting t-statistics, with statistical significance assessed via 4,000 permutation-based false discovery rate (FDR) corrections (q < 0.05) and 16 iterations of topology-informed pruning. Analyses were conducted separately for PACAP and cortisol and repeated at multiple T-thresholds (2.0, 2.5, 3.0), as done in prior studies [48, 49]. Lower t-score thresholds enhance sensitivity to subtle effects, while higher thresholds increase specificity. See the Supplement for additional details of the connectometry analysis.

### Statistical Analyses

For each white matter tract (UF, frontal parahippocampal cingulum, BNST–mPFC), and each hormone (cortisol, PACAP), associations between hormone levels and tract-average FA were examined using linear mixed-effects models implemented in the lme4 package in R [50]. Each model included tract-average FA from the left and right hemisphere as repeated outcome measurements and hormone level, hemisphere, age, sex, motion, assay batch, and draw time as fixed effects. Random intercepts were included to account for within-subject correlations between hemispheres. Interaction terms between hormone levels and hemisphere were tested in separate models to assess potential lateralized effects. Multiple-degree-of-freedom tests for regression parameters were performed to determine whether cortisol batch was a significant main effect using the *linearHypothesis* function from the car package. Extreme values (>3 standard deviations above the mean) were winsorized for cortisol (n=2) and PACAP (n=4), in line with prior work [11, 29, 33, 51]. For clarity in interpretation, unadjusted beta coefficients from all models were scaled by a factor of 1,000. No multiple testing correction was applied to results for tract-average FA; test-wise p-values are reported.

To assess segment-level microstructural associations, we conducted restricted connectometry analyses for each stress hormone as the regressor of interest (see above). We performed two sets of sensitivity analyses to evaluate the robustness of significant findings across the diffusion acquisition protocols: (1) repeating analyses with diffusion protocol as a nuisance regressor, and (2) repeating analyses after excluding participants scanned with the HCP Young Adult diffusion protocol. Results of these sensitivity analyses are presented in the Supplement. Finally, for streamlines identified as significantly associated with hormone levels at T = 3.0 with connectometry, we conducted post hoc linear regressions to examine associations between FA values and clinical symptom severity (CAPS-5 total, hyperarousal subscale, BDI-total scores), adjusting for age, sex, motion, draw time, and assay batch. Logistic regression was used to determine whether FA values were associated with PTSD diagnosis using the same covariates.

## RESULTS

### Circulating Cortisol and Tract-Average FA

In the UF model, circulating cortisol was not significantly associated with tract-average FA (β = −0.20, 95% CI [−1.00, 0.61], *p* = .63). Significant main effects emerged for left hemisphere (β = −10.61, 95% CI [−12.68, −8.5], *p* < .0001) and age (β =-0.51, 95% CI [−0.94, −0.08], *p* = 0.021), indicating higher FA in the right hemisphere and lower FA with increasing age. No other main effect reached statistical significance (p’s > .12).

In the frontal parahippocampal cingulum model, cortisol was not significantly associated with tract-average FA (β =-0.96, 95% CI [−2.12, 0.20], *p* =0.11). A significant left hemisphere effect was observed (β = - 39.10, 95% CI [−42.89 to −35.32], *p* < .0001) indicating higher FA in the right hemisphere, but no other main effects reached significance (*p*’s > .65).

In the BNST–mPFC model, cortisol was not significantly associated with tract-average FA (β = −1.06, 95% CI [−2.15, 0.06], *p* = .06). Significant main effects were found for left hemisphere (β = −55.20, 95% CI [−59.05, −51.35], *p* < .0001) and draw time (β = −0.08, 95% CI [−0.15, −0.007], *p* = .03), indicating higher FA in the right hemisphere. No other main effect reached statistical significance (*p*’s > 0.11).

No cortisol-by-hemisphere interaction was significant for any of the three tracts (Table S2).

### Circulating Cortisol and Localized White Matter FA

Restricted connectometry revealed significant negative associations between cortisol and white matter FA across all T-thresholds tested (see Figure 1; Table 2). At the lowest T-threshold, higher cortisol levels were associated with lower FA of streamlines associated with 14 white matter tracts. The strongest association, observed at the highest T-threshold (T = 3.0), revealed that higher circulating cortisol was significantly associated with lower FA of streamlines in the right dorsal cingulum. This cluster overlapped with the frontal-parietal, parolfactory, and frontal parahippocampal cingulum (FDR = .0018). No positive associations were found at any T-threshold. The negative association between cortisol and FA in the right dorsal cingulum remained significant after controlling for diffusion protocol (Figure S6) and in analyses limited to participants scanned with the HCP Adult Lifespan protocol (Figure S8).

**Figure 1.**
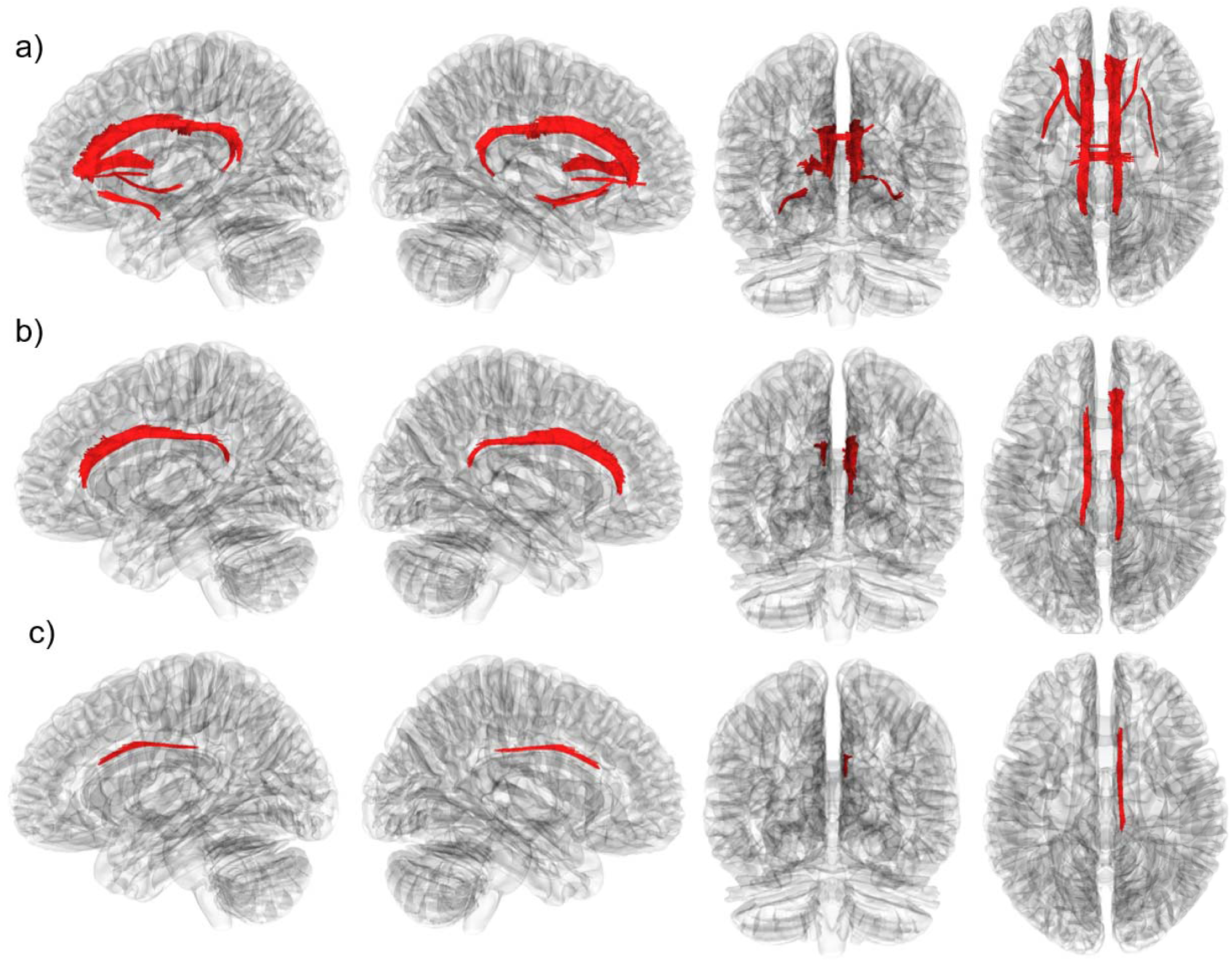
Restricted connectometry results of cortisol associations with white matter fractional anisotropy (FA). Panels show white matter segments (red) where higher cortisol levels correlated with lower FA at different T-thresholds: (a) T = 2.0; (b) T = 2.5; (c) T = 3.0.

**Table 2.** Results of restricted connectometry at each T-threshold where higher cortisol associated with lower white matter fractional anisotropy.

|  | **T = 2.0 threshold** |  |  | **T = 2.5 threshold** |  |  | **T = 3.0 threshold** |  |  |
| --- | --- | --- | --- | --- | --- | --- | --- | --- | --- |
| **Associated Tract Name** | Number of Tracts | Mean Length (mm) | FDR Value | Number of Tracts | Mean Length (mm) | FDR Value | Number of Tracts | Mean Length (mm) | FDR Value |
| Right Frontal Parietal Cingulum (association) | 2113 | 40.10 | <.0001 | 668 | 37.90 | < .001 | 86 | 36.86 | =.002 |
| Right Parolfactory Cingulum (association) | 940 | 44.97 | <.0001 | 348 | 42.86 | < .001 | 34 | 39.84 | =.002 |
| Right Frontal Parahippocampal Cingulum (association) | 484 | 40.55 | <.0001 | 161 | 36.90 | < .001 | 26 | 32.85 | =.002 |
| Left Frontal Parahippocampal Cingulum (association) | 735 | 38.03 | <.0001 | 126 | 34.57 | < .001 |  |  |  |
| Left Parolfactory Cingulum (association) | 215 | 34.83 | <.0001 | 1 | 32.02 | < .001 |  |  |  |
| Left Frontal Parietal Cingulum (association) | 126 | 36.45 | <.0001 | 48 | 33.92 | < .001 |  |  |  |
| Left Anterior Thalamic radiation (basal ganglia projection) | 284 | 33.51 | <.0001 |  |  |  |  |  |  |
| Body of Corpus Callosum (commissure) | 41 | 32.44 | <.0001 |  |  |  |  |  |  |
| Left Uncinate Fasciculus (association) | 32 | 32.75 | <.0001 |  |  |  |  |  |  |
| Right Inferior Fronto-Occipital Fasciculus (association) | 20 | 34.81 | <.0001 |  |  |  |  |  |  |
| Right Anterior Thalamic Radiation (basal ganglia projection) | 19 | 33.90 | <.0001 |  |  |  |  |  |  |
| Left Anterior Corticostriatal Tract (basal ganglia projection) | 13 | 31.54 | <.0001 |  |  |  |  |  |  |
| Left Inferior Fronto-Occipital Fasciculus (association) | 12 | 32.50 | <.0001 |  |  |  |  |  |  |

Note: parenthetic text list the type of broader white matter tract. FDR: false discovery rate.

### Circulating PACAP and Tract-Average FA

PACAP was not significantly associated with tract-average FA in any of the three tracts. In the UF, the association was non-significant (β = −0.0005, 95% CI [−0.006, 0.005], *p* = .86), although significant effects were observed for left hemisphere (β = −10.91, 95% CI [−13.08, −8.74], *p* < .0001), age (β = −0.56, 95% CI [−0.99, −0.14], *p* = 0.01), and PACAP batch (β = −9.55, 95% CI [−18.36, −0.75], *p* =.03). No other main effect reached statistical significance (*p*’s > 0.08).

Similarly, PACAP was not associated with tract-average FA of the frontal parahippocampal cingulum (β = 0.005, 95% CI [−0.003, 0.01], p = .22), though a significant effect of left hemisphere was observed (β = - 38.58, 95% CI [−42.53, −34.63], *p* < .0001); all other covariates were non-significant (*p*’s > 0.71).

In the BNST–mPFC model, PACAP was also not significantly associated with tract-average FA (β = 0.002, 95% CI [−0.006, 0.009], *p* = .67), but significant effects were observed for left hemisphere (β = −55.48, 95% CI [−59.53, −51.44], *p* < .0001) and PACAP batch (β = −12.73, 95% CI [−24.91, −0.55], *p* = .04). No other main effect reached statistical significance (*p*’s > 0.21).

No PACAP-by-hemisphere interaction was significant for any of the three tracts (Table S3).

### Circulating PACAP and Localized White Matter FA

As shown in Figure 2 and Table 3, restricted connectometry identified significant associations between PACAP and white matter FA across all T-thresholds. At the lowest T-threshold, higher PACAP levels were associated with lower FA in streamlines from five white matter tracts, including the inferior longitudinal fasciculus, corpus callosum, and hippocampal alveus. However, these findings did not persist at higher T-thresholds.

**Figure 2.**
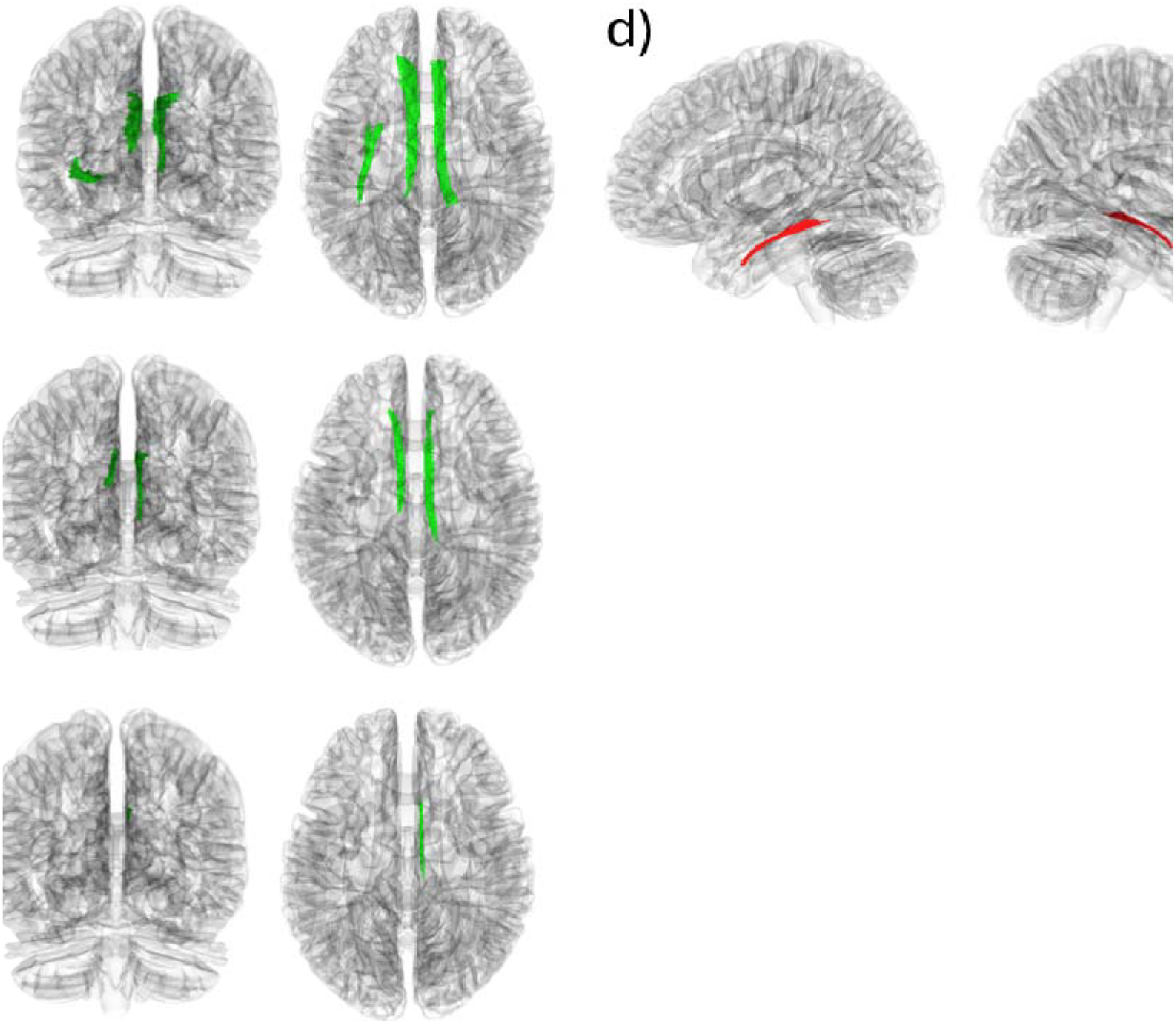
Restricted connectometry results of PACAP associations with white matter fractional anisotropy (FA). Panels (a-c) show white matter segments (green) where higher PACAP levels correlated with higher white matter FA at different T-thresholds: (a) T = 2.0. (b) T = 2.5; (c) T = 3.0. Panel (d) shows white matter segments (red) where higher PACAP levels correlated with lower white matter FA at T = 2.0, none remaining significant at higher T-thresholds.

**Table 3.**
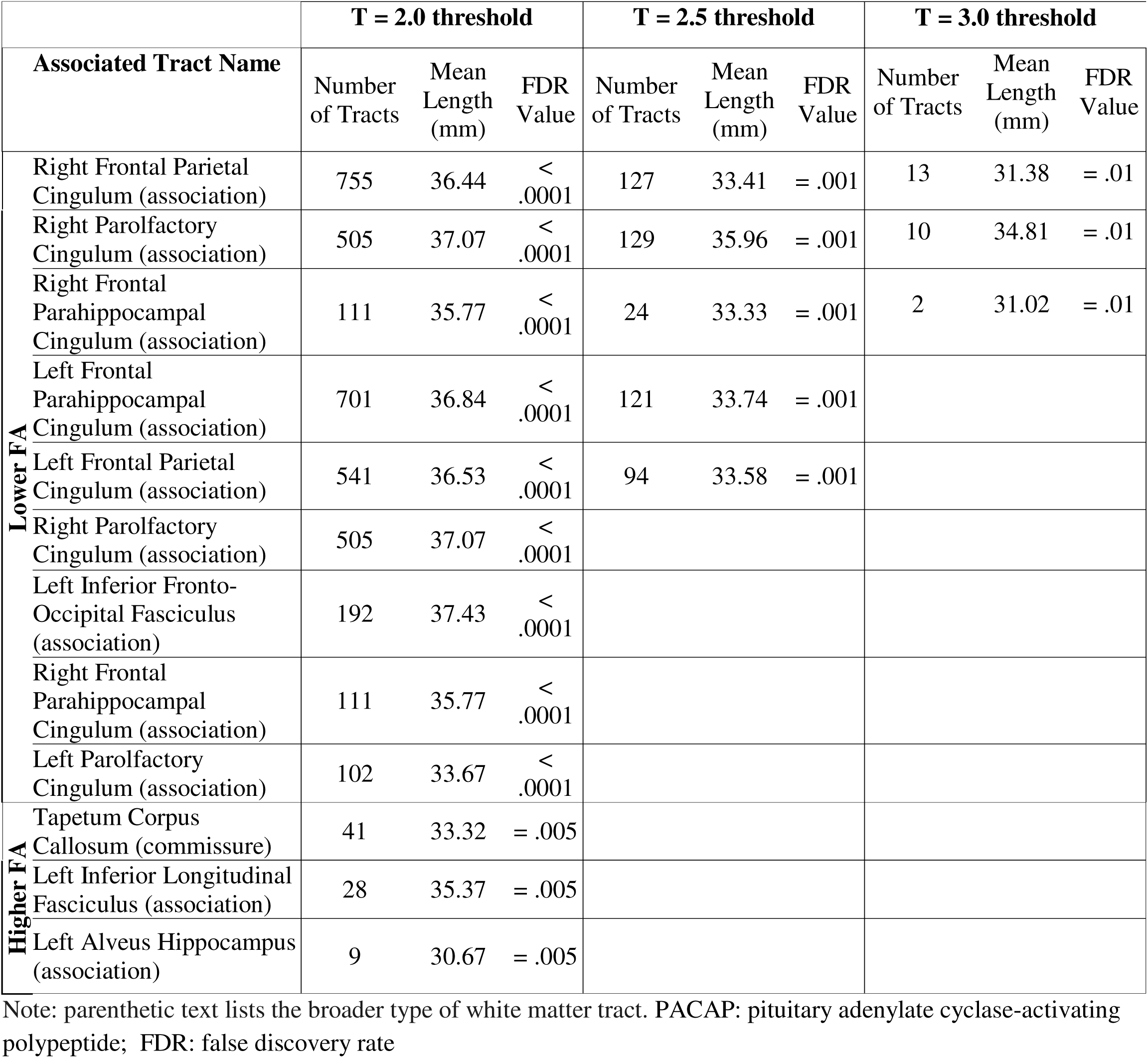
Results of restricted connectometry at each T-threshold where higher PACAP is associated with higher or lower white matter fractional anisotropy.

Conversely, higher PACAP was associated with higher FA in streamlines corresponding to seven white matter tracts, including portions of the cingulum and inferior frontal-occipital fasciculus. The strongest finding, observed at the highest T-threshold (T = 3.0), showed that higher PACAP was associated with higher FA in the right dorsal cingulum, overlapping with frontal-parietal, parolfactory, and frontal parahippocampal cingulum streamlines (*FDR* = .012). This association remained significant after controlling for diffusion protocol (Figure S7) and when restricted to participants scanned with the HCP Adult Lifespan protocol (Figure S9).

### Clinical Symptom Associations with Stress Hormone-Linked FA segments

Post hoc analyses examined whether FA of streamlines significantly associated with cortisol or PACAP at T = 3.0 were related to clinical symptom severity. For cortisol-associated streamlines, FA was not associated with either CAPS-5 total (β = −14.81, *p* = .12), CAPS-5 hyperarousal severity (β = −3.71, *p* = .27), BDI-II total score (β = −13.38, *p* = .16), or PTSD diagnosis (β = −0.16, *p* = .94), controlling for age, sex, motion, draw time, and assay batch. For PACAP-associated streamlines, no significant associations were observed with either CAPS-5 total (β = −13.38, *p* = 0.16), hyperarousal severity (β = −4.64, *p* = .17), BDI-II total scores (β = −12.83, *p* = .16), or PTSD diagnosis (β = −0.07, *p* = .97) controlling for the same covariates. Cortisol levels were not significantly correlated with PACAP levels (Pearson’s correlation coefficient = 0.09, *p* = .32).

## DISCUSSION

This investigation advances our understanding how peripheral stress hormones relate to white matter microstructure in PTSD. While no associations were observed between cortisol or PACAP and tract-average microstructural integrity of the UF, frontal parahippocampal cingulum, or BNST–mPFC tracts, restricted connectometry revealed distinct and spatially localized associations in the dorsal cingulum. Specifically, higher circulating PACAP was associated with higher FA, whereas higher cortisol was linked to lower FA in this region. These associations were not related to PTSD diagnosis, total PTSD symptoms, hyperarousal, or depression symptom severity, suggesting that they may reflect hormone-specific biological pathways – perhaps biological subtypes – rather than general symptom or stress burden. To our knowledge, this is the first study to demonstrate these opposite associations in PTSD, offering a potential explanation to reconcile previously inconsistent findings on cingulum microstructure in the disorder.

These findings emerge amongst mixed findings on dorsal and anterior cingulum microstructural integrity in PTSD literature (reviewed in [52, 53]). Some studies have reported higher dorsal cingulum FA in persistently symptomatic versus remitted PTSD patients [54, 55]. Conversely, others have found lower cingulum FA in PTSD patients. For example, PTSD patients show lower FA in the left dorsal segment compared to healthy subjects [56], in the right anterior segment compared to those with generalized anxiety disorder [57], and lower FA of the dorsal cingulum in cases of PTSD comorbid with alcohol use disorder compared to alcohol use alone [20]. Our results suggest that individual differences in cortisol and PACAP levels may contribute to these inconsistent findings, offering biologically plausible mechanisms underlying heterogeneity in dorsal cingulum microstructure in PTSD.

Cortisol and PACAP engage distinct yet partly overlapping biological pathways that may differentially affect white matter integrity. Elevated peripheral cortisol, a steroid-based stress hormone, is pro-inflammatory [58] and can impair myelination [59]; peripheral inflammatory markers have been linked to lower anisotropy in the cingulum bundle [60]. Notably, cortisol is dynamically regulated. While PTSD has been associated with suppression of cortisol levels in some studies [61–63], major depression has been more frequently been associated with non-suppression following HPA stimulation [64]. In contrast, [61–64]PACAP – a peptide stress hormone – is generally considered anti-inflammatory [65–69], although it may promote pro-inflammatory signaling under certain conditions, potentially depending on receptor subtype expression (PAC1, VPAC1, and VPAC2) [70]. PACAP has also been shown to promote neurotrophic processes, including axonal outgrowth [71], synaptic plasticity [72], and growth factor expression [73], all of which may support white matter integrity [74]. The PAC1 receptor, which has the highest affinity for PACAP [75] and is implicated in PTSD [5], mediates many of PACAP’s anti-inflammatory and neuroprotective effects, including pro-inflammatory cytokine suppression [65, 69]. Together, these divergent biological functions raise the possibility that PACAP and cortisol exert opposing effects on white matter microstructure. Such a framework could help explain why higher PACAP levels in our study were associated with greater dorsal cingulum integrity, whereas higher cortisol levels correlated with lower integrity. Further research is needed to test this hypothesis directly.

The regional specificity of our findings may have implications for PTSD-related behavioral phenotypes. The dorsal (anterior and mid) cingulum supports affective functions, including fear, reward processing, and emotional regulation [76]. In contrast, the posterior or parahippocampal cingulum is more involved in cognitive functions such as cognitive control and spatial memory [52, 76]. Our findings suggest that PACAP and cortisol may exert differential effects on emotional processes in PTSD through localized influences on dorsal cingulum microstructure. Although FA in this region was not significantly associated with clinical symptom scores, it may relate to behavioral domains not well captured by standard clinical measures.

One such domain is sleep, which is frequently disrupted in PTSD and influenced by stress hormones. Rodent studies have shown that CRF and PACAP affect sleep architecture differently, with infusion of anxiogenic doses of PACAP decreasing wakefulness and increasing slow wave and rapid eye movement sleep [10]. In contrast, infusion of anxiogenic doses of CRF causes more transient increases in slow wave sleep during certain phases of the diurnal cycle. In non-PTSD samples, sleep disturbances have been associated with dorsal cingulum white matter alterations [77–81] and with whole-brain white matter alterations in PTSD [82]. These lines of evidence highlight the need for further investigation into whether stress hormones differentially influence sleep via dorsal cingulum pathways in PTSD.

A key strength of this study is the use of region-restricted connectometry, which enabled detection of localized FA associations not evident in tract-average analyses. This approach aligns with emerging literature emphasizing along-tract and segment-level diffusion analyses to capture anatomically focal effects [83–87]. Unlike along-tract analysis that requires averaging across white matter bundles, connectometry identifies specific white matter areas influenced by high or low FA, preserving spatial specificity. Connectometry has previously been used at the whole-brain level [88–91], including in psychiatric and trauma-exposed cohorts [92] [49], and to identify white matter correlates of peripheral inflammation [93]. To our knowledge, our study is the first to restrict connectometry based on *a priori* hypotheses, enabling the identification of white matter associations relevant to areas of interest without averaging.

Nonetheless, several limitations should be noted. First, the literature on structural characterization of the BNST–mPFC white matter pathway is scarce, and our deterministic tractography approach, while informed by anatomical studies [94], requires validation with higher-resolution or histologically guided imaging [95]. Second, while our connectometry was constrained to the UF, frontal parahippocampal cingulum, and BNST– mPFC, some streamlines extended into adjacent regions. Cortisol was associated with lower FA in the anterior thalamic radiation, corpus callosum, inferior frontal-occipital fasciculus, and anterior corticostriatal tract. PACAP was associated with higher FA in the inferior frontal-occipital fasciculus and lower FA of the tapetum of the corpus callosum, inferior longitudinal fasciculus, and alveus of the hippocampus. These findings appeared only at lower T-thresholds and did not persist at more stringent levels, suggesting they may be spurious. Furthermore, because cortisol and PACAP levels were assessed at a single morning time point, we could not capture diurnal variability or dynamic responses to stress. Finally, connectometry allow streamlines to pass through or end in a specific area, but not to be entirely confined to a given ROI, introducing interpretative challenges. We opted for a “passing-through” mask because an “ending” criterion introduced more off-target streamlines, but we acknowledge this trade-off.

An additional interpretive challenge stems from the complex fiber architecture of the dorsal cingulum, where multiple tracts overlap, including the frontal-parietal, parolfactory, and frontal parahippocampal cingulum. Our highest-threshold findings fell within this complex area. Thus, we interpret our results as implicating the broader dorsal cingulum and caution against over-assigning findings to specific cingulum subdivisions (see Supplement, Figure S10).

In conclusion, this study provides novel evidence that peripheral fasting PACAP and cortisol levels are differentially associated with dorsal cingulum microstructure in adults with posttraumatic stress. These findings highlight the importance of investigating the nuanced and regionally specific correlates of stress hormones in PTSD neurobiology and potentially reconcile inconsistencies in prior white matter findings. Future research should examine how stress hormones relate to dorsal-cingulum-supported behavioral functions, such as sleep and arousal regulation, and apply regionally targeted diffusion analyses to refine neurobiological models of PTSD.

## Data availability

Data are available through the NIMH National Data Archive

(NDA; https://nda.nih.gov/edit_collection.html?id=3166), and upon reasonable request to the senior author, IMR.

## Acknowledgements

We are deeply grateful to the individuals who participated in this study. Their generosity in sharing their time and experiences made this research possible and contributes meaningfully to advancing understanding of posttraumatic stress. We also thank the MRI Technologists of the McLean Imaging Center

## Author Contributions

**SJG**: Methodology, Software, Validation, Formal analysis, Data curation, Writing – original draft, Writing – review & editing, Visualization. **SAJ**: Methodology, Formal analysis, Investigation, Data curation, Writing – original draft, Writing – review & editing, Visualization. **CR**: Methodology, Formal analysis, Writing – review & editing. **QD**: Methodology, Writing – review & editing. **EA**: Methodology, Data curation, Investigation, Writing – review & editing. **JUB**: Methodology, Writing – review & editing. **VM**: Resources, Writing – review & editing. **SEH**: Resources, Writing – review & editing. **WAC**: Funding acquisition, Writing – review & editing. **KJR**: Conceptualization, Writing – review & editing, Funding acquisition. **SLR**: Conceptualization, Supervision, Writing – review & editing, Funding acquisition. **IMR**: Conceptualization, Methodology, Validation, Formal analysis, Investigation, Resources, Writing – original draft, Writing – review & editing, Supervision, Project administration, Funding acquisition.

## Funding

This work was supported by NIH award P50MH115874 (to WAC and KJR, PDs; IMR, SLR, Project 4 PIs. CR). In addition, the investigators were partially supported by R01MH120400 (IMR), R01MH125852 (IMR), R01MH097988 (SEH and VM).

## Competing Interests

Within the last 3 years WAC has served as a Consultant for AbbVie, Neumora, and Psy Therapeutics, and has Sponsored Research Agreements from AbbVie and Delix. No funding from these entities was used to support the current work. SLR is employed by Mass General Brigham/McLean Hospital, has received payment as secretary of Society of Biological Psychiatry, and for Board service to Mindpath Health/Community Psychiatry and National Association of Behavioral Healthcare, served as volunteer member of the Board for The National Network of Depression Centers, received royalties from Oxford University Press, American Psychiatric Publishing Inc, and Springer Publishing, and has received research funding from NIMH. Dr. Ressler has performed scientific consultation for Bioxcel, Bionomics, Acer, Leal Therapeutics, and Jazz Pharma; serves on Scientific Advisory Boards for Sage, Boehringer Ingelheim, Senseye, and the Brain Research Foundation, and he has received sponsored research support from Alto Neuroscience. He has received research funding from the NIH, DARPA, DOD and the Wellcome Leap Foundation.

## SUPPLEMENTAL MATERIALS

### MATERIALS AND METHODS

#### Participants

Exclusion criteria were left-handedness; medical conditions that could confound results (e.g., untreated seizure disorder); inability to tolerate blood draws; history of head trauma with loss of consciousness > 5 minutes; current treatment with an antipsychotic (unless prescribed for PTSD-related sleep disturbances); past month moderate-to-severe alcohol use disorder or substance use disorder; current psychotic disorder, anorexia nervosa, obsessive-compulsive disorder, manic or mixed mood episode; lifetime history of schizophrenia or schizoaffective disorder; MRI contraindications (e.g., metal implants, claustrophobia); positive pregnancy test on the day of scanning for female participants; and history of hormonal replacement therapy or surgery to change biological sex.

#### MRI Data Acquisition

For data acquired with the HCP Young Adult protocol, a T1-weighted 3D MPRAGE sequence was acquired (TR =2400 ms, TE = 2.14 ms; flip angle = 8 deg, FOV = 224 × 224, voxel size = 0.7 mm isotropic), along with a high-angular multi-shell whole brain diffusion-weighted imaging sequence (TR = 5520, TE = 89.5 ms; flip angle = 78 deg, refocusing flip angle = 160 deg, FOV = 210 × 180; matrix = 168 × 144, slice thickness = 1.25 mm, 111 slices, voxel size = 1.25 mm isotropic, b = 1000, 2000, and 3000 s/mm2, _∼_30 directions/shell, multiband factor = 3). For data acquired with the HCP Adult Lifespan protocol, a T1-weighted MPRAGE sequence was acquired (TR = 2500 ms, TE = 1.81/3.6/5.39/7.18 ms, flip angle = 8 deg, FOV = 256 × 240, voxel size = 0.8 mm isotropic), along with a high-angular multi-shell whole brain diffusion-weighted imaging sequence (TR = 3230, TE = 89.5 ms; flip angle = 78 deg, refocusing flip angle = 160 deg, FOV = 210 × 210; matrix = 168 × 144, slice thickness = 1.5 mm, 92 slices, voxel size = 1.5 mm isotropic, b = 1500 and 3000 s/mm2, _∼_47 directions/shell, multiband factor = 4).

#### Diffusion Imaging Processing

Additional preprocessing included the *–repol* option, which was used to detect and replace outliers, further improving data quality [1]. Participant motion was quantified with FSLs’ *eddy_quad* quality control tool, which calculates average motion relative to the preceding diffusion-weighted image volume; motion estimates were included as nuisance regressors in statistical models [2]. Specifically, participants with an *R*^2^ value less than 0.6 were flagged, consistent with prior studies [3–7]. All flagged scans underwent visual inspection. Of the 9 individuals excluded due to image quality reasons: six participants were excluded due to poor image registration; one due to signal dropout in the prefrontal cortex; another due to poor template fit identified during visual inspection; and lastly, one final participant was excluded due to low (lower than 3 standard deviations below the sample mean) neighboring diffusion-weighted imaging correlation (NDC). NDC, a quality-control measure derived from DSI Studio, assesses the consistency of diffusion signals between neighboring volumes; low NDC values indicate low data quality, potentially due to residual eddy current artifacts, excessive head motion, or hardware-related issues [8–10].

#### BNST**–**mPFC Custom Tractography

To develop BNST–mPFC tractography, we first created a population-average BNST mask by combining images from manually segmented BNST regions across 123 participants’ T1-weighted images. These images were processed using Freesurfer 6 [11] skull-stripped using AFNIs 3dSkullStrip [12] and corrected for signal intensity. Manual BNST segmentation was conducted according to methods outlined by Theiss et al. (2017). Briefly, the BNST was traced in the coronal view with the superior aspect of the anterior commissure serving as the inferior boundary. The superior boundary was defined by either the thalamostriate vein or ventral aspect of the caudate body. The medial and lateral boundaries were defined by the fornix and internal capsule, respectively. The anterior boundary was defined by two slices or 1.8mm anterior to the most anterior slice in which the anterior commissure is still visibly joined at the midline. To create the population-average mask, we averaged all manually segmented masks and removed voxels that were categorized as BNST in less than 10% of participants. The resulting BNST masks were then used as “end” regions for tractography. Previous research has identified an anterior BNST pathway that traverses the transition area of the nucleus accumbens and caudate nucleus and enters the prefrontal white matter *en route* to the mPFC and orbitofrontal cortex [14]. In detailed anatomical work, Kamali et al. (2023) have identified this pathway as a part of the prefronto-caudo-thalamic tract. This white matter pathway has previously been delineated using a vertical coronal plane as a “region-of-interest” (ROI) placed at the head of the caudate nucleus and passing through the intersection of the genu and rostrum of the corpus callosum [16].

Following this precedent, we created a vertical coronal plane (MNI coordinate bounding box: (x = - 244, y = −76, z = −14) to (x = 2, y = −76, z = 260)) to guide tractography located twelve coronal slices posterior to the most anterior part of the genu of the corpus collosum. This plane acted as an ROI (shown in **Figure S1)**, thereby capturing white matter streamlines that ended in the BNST and passed to the mPFC. This approach differentiates BNST–mPFC pathways from adjacent tracts, including the anterior thalamic radiation [16]. Visualization of BNST–mPFC streamlines against the anterior thalamic radiation (derived using augmented tractography) from our data are shown in **Figure S2**. Additional tractography settings included an anisotropy threshold that was randomly selected between 0.5 and 0.7 otsu threshold, an angular threshold of 65 degrees, the step size was set to voxel spacing, streamlines with length shorter than 0.0 or longer than 200.0 mm were discarded and a total of 5,000,000 seeds were placed. Tractography was conducted in two iterations, once using the left BNST population-level mask as the “end” region and once using the right BNST population-level mask. Both iterations used the coronal ROI previously described. The resulting streamlines closely resemble those reported in Krüger et al. 2015, supporting the validity of our tractography approach and its ability to delineate this white matter pathway. Images of the BNST region, coronal ROI, and BNST–mPFC streamlines are shown in **Figure S3**.

#### Uncinate Fasciculus and Frontal Parahippocampal Cingulum Tractography

For UF and frontal parahippocampal cingulum tracts, we used augmented tractography. Augmented tractography incorporates three strategies to enhance reproducibility: parameter saturation, automatic atlas-based tract recognition/filtering, and topology-informed pruning. Parameter saturation involves random combinations of anisotropy thresholds, step sizes, and angular thresholds to saturate the parameter space, generating multiple tracking iterations with unique parameters. Each iteration is then filtered with automatic atlas-based filtering, which selects for the shortest Hausdorff distance with the trajectories of the HCP842 tractography atlas [17]. Topology-informed pruning consists of identifying singular or low tract density streamlines as false connections for removal [10].

#### Restricted Connectometry Analyses

To constrain the analysis to our tracts of interest, we converted the bilateral UF, frontal parahippocampal cingulum, and BNST–mPFC streamlines into a single binary mask (**Figure S5**). This mask was then used as a ROI to detect localized white matter segments with FA values that correlate with cortisol. For complete description of connectometry analysis see the Supplement. This procedure involved first computing the linear fitting between all covariates (including cortisol) and FA of each voxel. Then, for each covariate excluding cortisol and the intercept, FA values were adjusted using the coefficients computed from linear fitting. We then calculated the rank-based nonparametric Spearman correlation between cortisol and the adjusted FA measure. The resulting t-statistics were then used for subsequent deterministic fiber tracking [18]. Once correlational tractography was conducted, the statistical significance was evaluated using a permutation-based approach [19], with 4,000 randomized permutations of the group labels to generate the null distribution of tract lengths. To reduce false-positive streamlines, results were filtered using topology-informed pruning with 16 iterations [20]. These procedures are detailed in full in the method resource publication [19]. We applied varying t-score thresholds (T = 2.0, 2.5, 3.0) as done in prior studies [21,22]. Lower t-score thresholds enhance detection of subtle effects (high sensitivity), while higher t-score thresholds increase specificity thereby providing an overview of significant findings. Significant streamlines were identified using a false discovery rate (FDR) correction at q < 0.05. These analyses were repeated using PACAP as the regressor of interest.

### DISCUSSION

#### Considerations for Low T-Score Thresholds in Connectometry

An important consideration in interpreting our findings is that our strongest associations at T = 3.0 were located in portions of dorsal cingulum white matter that extend beyond the frontal-parahippocampal cingulum, such as the frontal parietal and parolfactory cingulum. These designations are based on the findings of a clustering algorithm that attempts to resolve overlapping white matter regions. Estimates suggest that approximately 90% of the brain’s white matter contains multiple or crossing fiber populations [23], and the dorsal cingulum is one such region, containing streamlines from several cingulum subdivisions.

The cortisol and PACAP-associated streamline identified in our findings fall within this complex area of multiple fibers, containing streamlines from the parolfactory, frontal parietal, and frontal parahippocampal tracts (**Figure S10)**. This anatomical complexity introducdes inherent challenge in assigning a singular, standardized white matter label to a small group of streamlines that may fall in an area of multiple crossing fibers belonging to a variety of possible cingulum subdivisions.

Therefore, we suggest caution when interpreting associations identified at lower T-score thresholds, as these may reflect adjacent and overlapping regions rather than our precise area of interest. While statistically significant, these associations were weaker, not persisting at higher T-score thresholds. In addition, we suggest caution in interpreting cingulum associations at the highest T-threshold as belonging to separate and specific cingulum subdivisions. However, we are confident that the observed cortisol and PACAP associations are colocalized within the broader dorsal cingulum region, even if they cannot be definitively ascribed to individual subcomponents.

### SUPPLEMENTAL FIGURES

**Figure S1.**
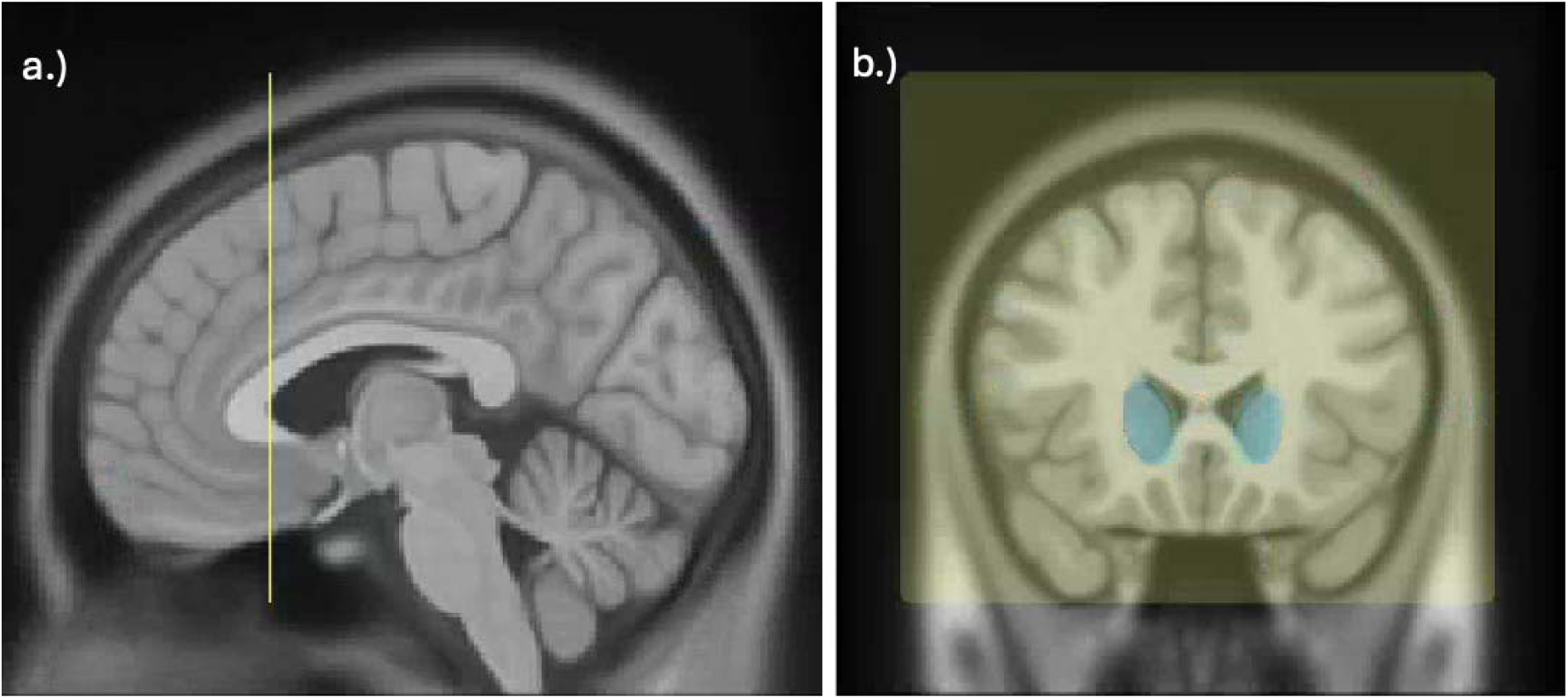
Region of interest (ROI) used to guide deterministic tractography of BNST–mPFC streamlines, analogous to that of Kamali et al [16]. (a) Sagittal midline view of the ROI (yellow) displaying the ROI positioned between the rostrum and genu of the corpus callosum. (b) Coronal view of the ROI (yellow), with the head of the caudate nucleus (blue) shown as a point of reference. *Note*: BNST: bed nucleus of the stria terminalis; mPFC: medial prefrontal cortex.

**Figure S2.**
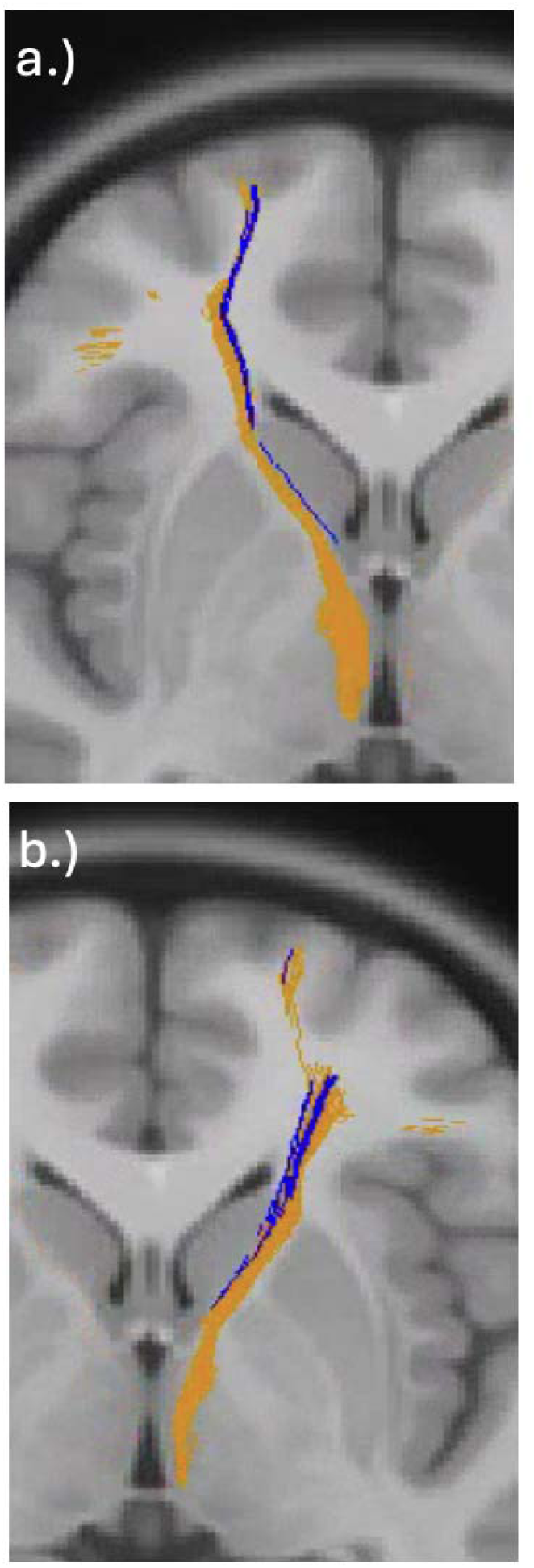
Visualization of BNST–mPFC streamlines dissociated from the anterior thalamic radiation (ATR). (a) Left BNST–mPFC tract (blue) traversing medially alongside the ATR (orange). (b) Right BNST–mPFC pathway (blue) traversing medially alongside the ATR (orange). Both images show BNST–mPFC pathways joining the anterior thalamic streamlines in the prefrontal white matter consistent with Kamali et al [16]. *Note*: BNST: bed nucleus of the stria terminalis; mPFC: medial prefrontal cortex.

**Figure S3.**
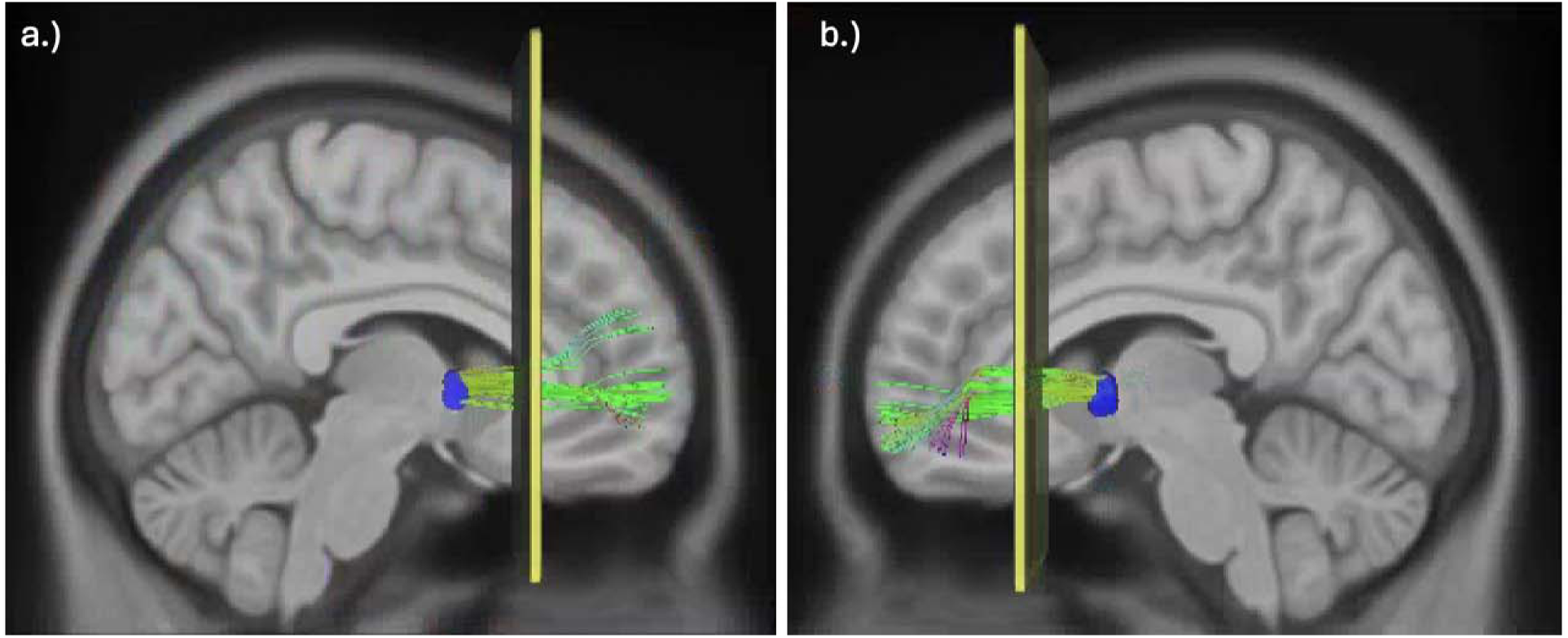
Deterministic tractography of left and right BNST–mPFC white matter streamlines. The BNST (blue) was used as an “end” region, and a coronal region-of-interest (yellow) was used to guide deterministic tractography. *Note*: BNST: bed nucleus of the stria terminalis; mPFC: medial prefrontal cortex.

**Figure S4.**
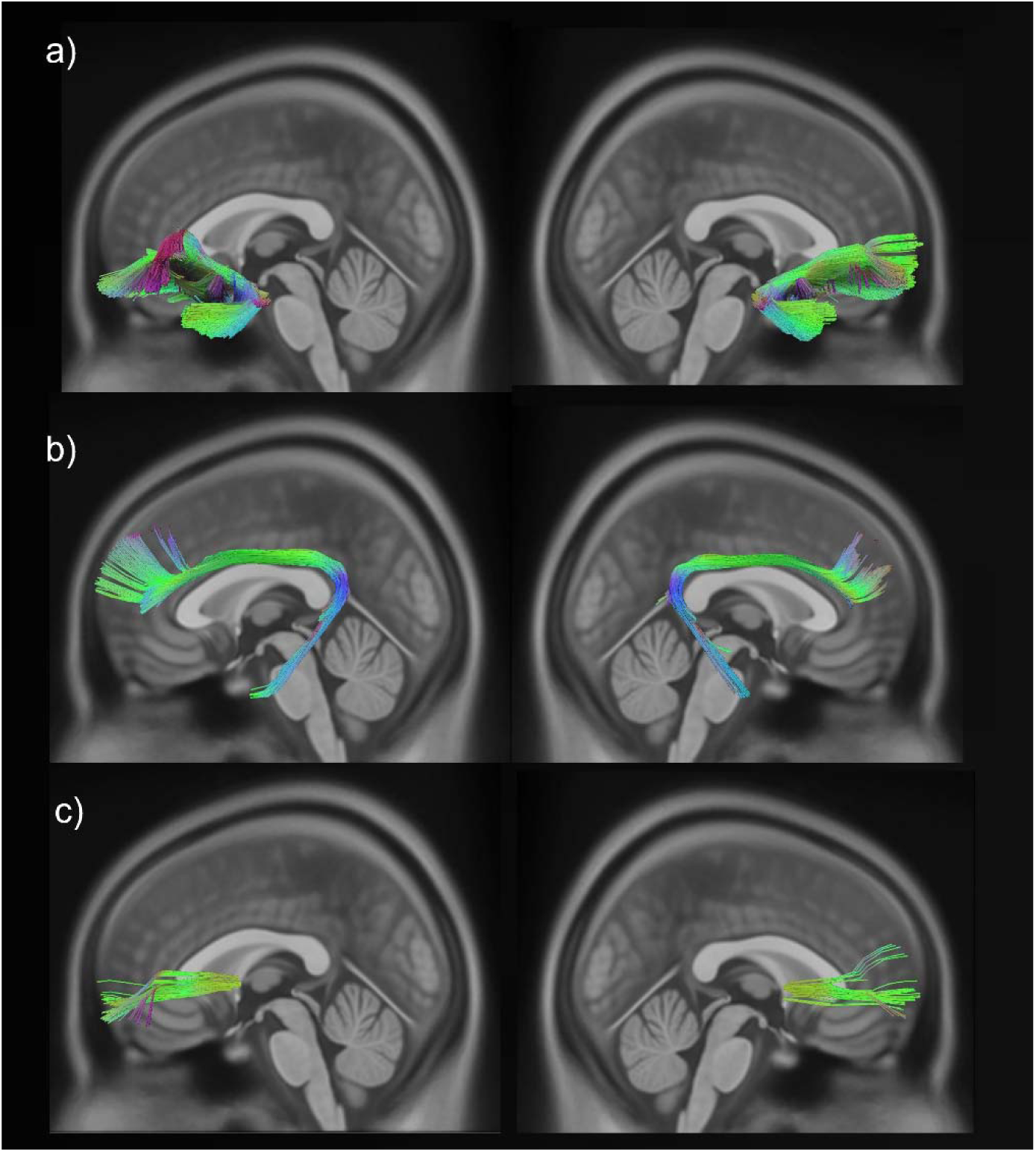
White matter tracts used for tract-average analyses. (a) Left and right uncinate fasciculus. (b) Left and right frontal parahippocampal cingulum. (c) Left and right BNST–mPFC. *Note*: BNST: bed nuclei of the stria terminalis; mPFC: medial prefrontal cortex.

**Figure S5.**
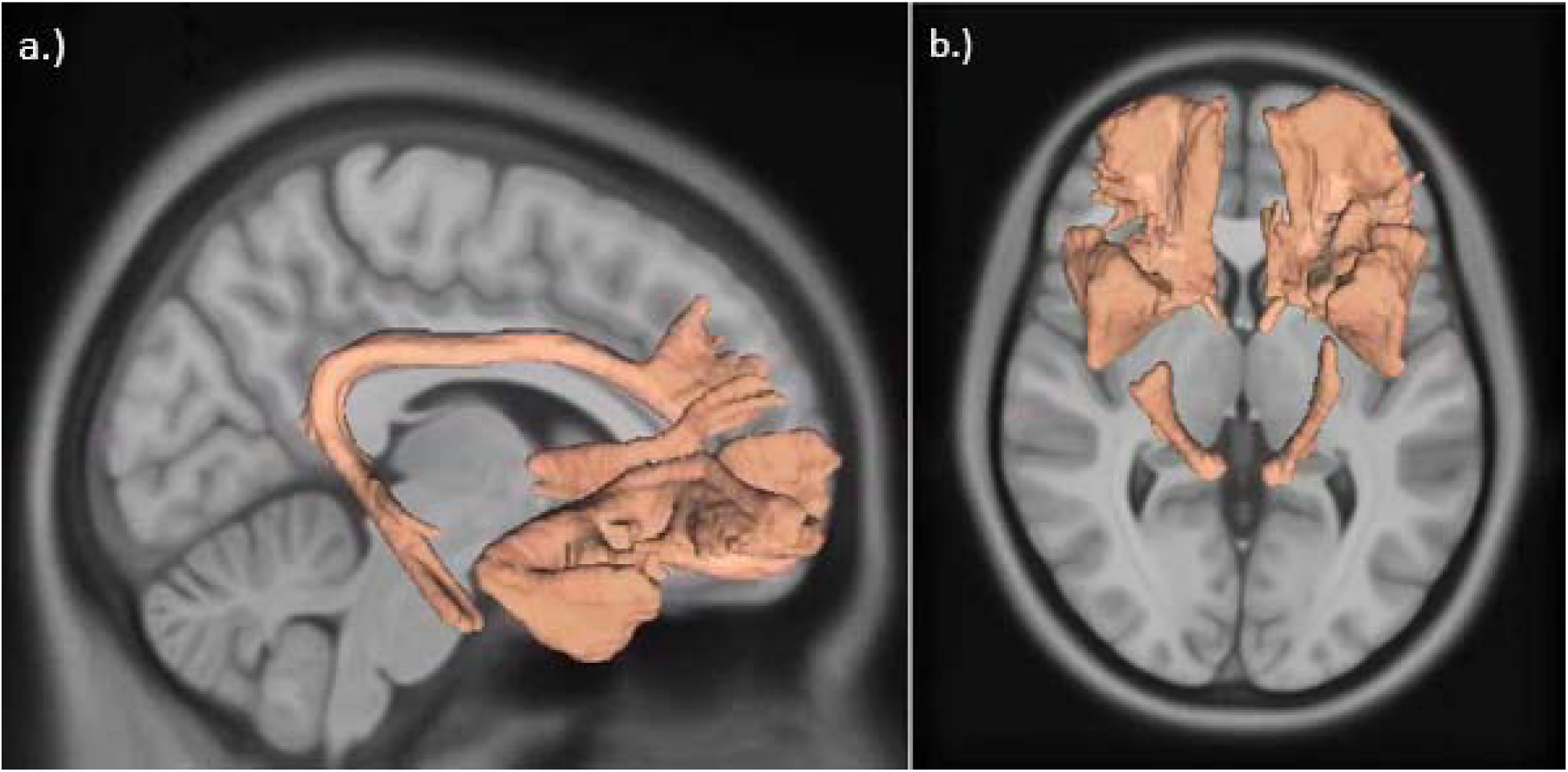
Binary mask used for restricted connectometry analyses. (a) mid-sagittal view. (b) axial view.

**Figure S6.**
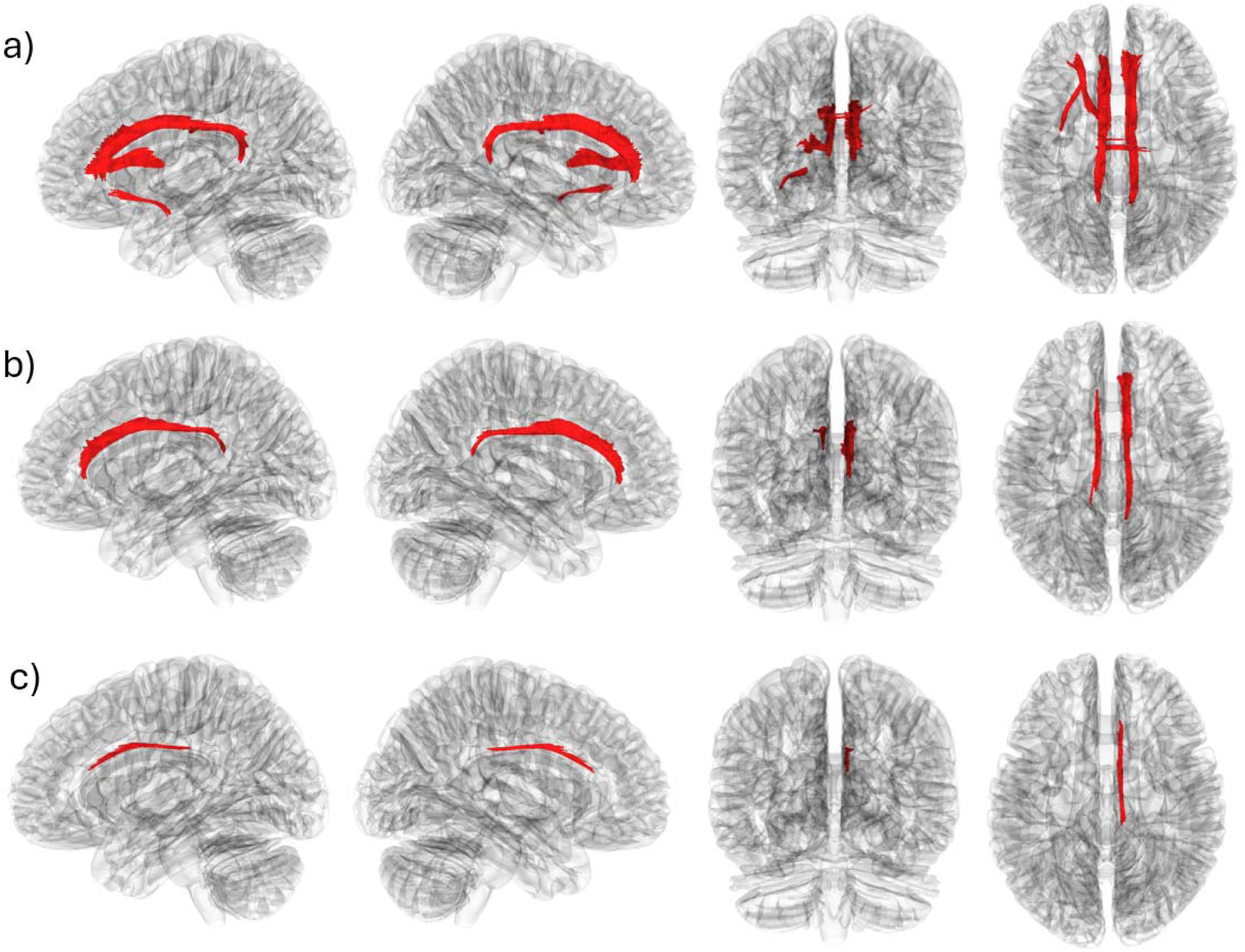
Restricted connectometry results of cortisol associations with white matter fractional anisotropy (FA), with diffusion protocol included as a covariate. All panels show white matter segments (red) where higher cortisol levels correlated with lower FA at different T-thresholds: (a) T = 2.0; (b) T = 2.5; (c) T = 3.0.

**Figure S7.**
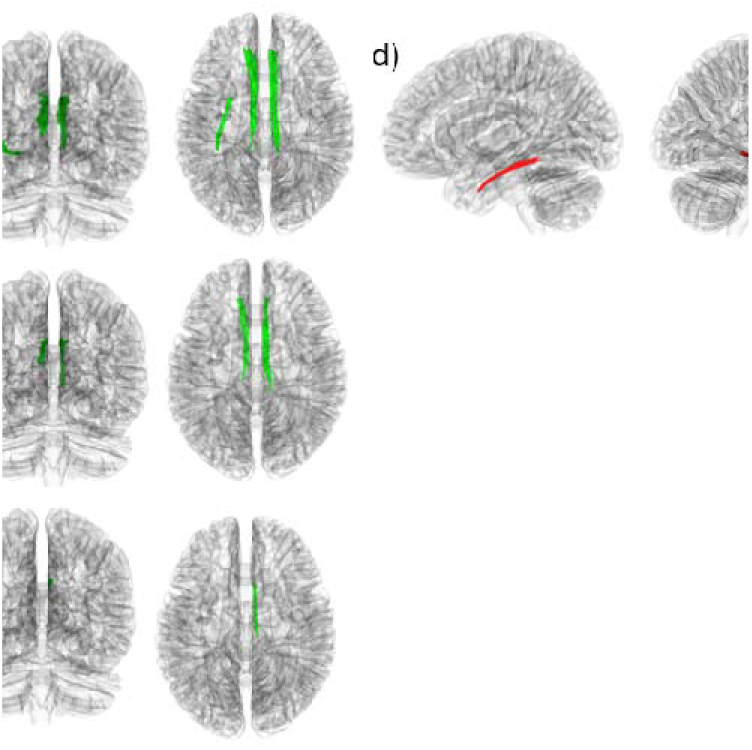
Restricted connectometry results of PACAP associations with white matter fractional anisotropy (FA), with diffusion protocol included as a covariate. Panels (a-c) show white matter segments (green) where higher PACAP levels correlated with higher white matter FA at different T-thresholds: (a) T = 2.0; (b) T = 2.5; (c) T = 3.0. Panel (d) shows white matter segments (red) where higher PACAP levels correlated with lower FA at T = 2.0, none remaining significant at higher T-thresholds.

**Figure S8.**
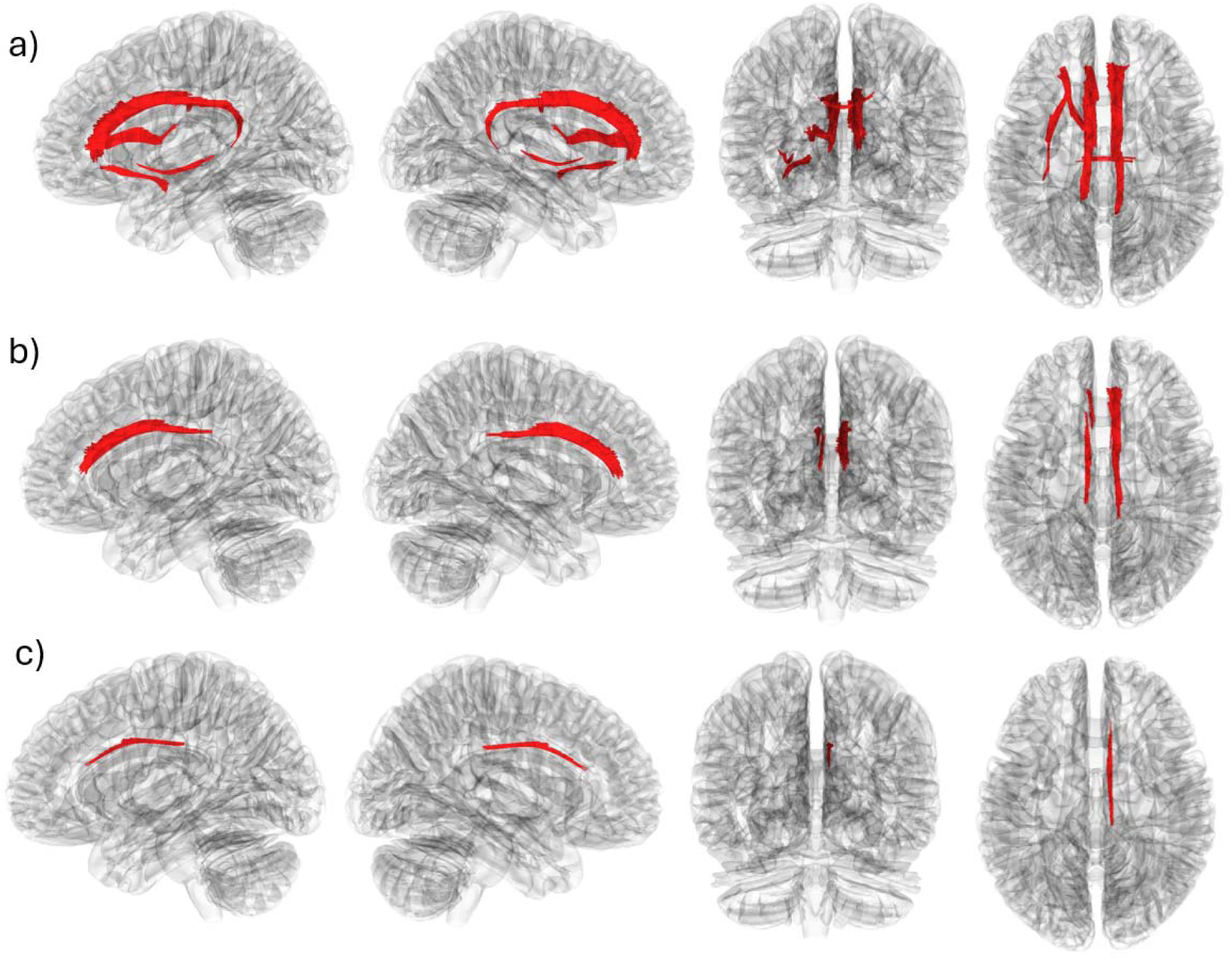
Restricted connectometry results of cortisol associations with white matter fractional anisotropy (FA), within participants scanned with the Adult Lifespan Protocol. All panels show white matter segments (red) where higher cortisol levels correlated with lower FA at different T-thresholds: (a) T = 2.0; (b) T = 2.5; (c) T = 3.0.

**Figure S9.**
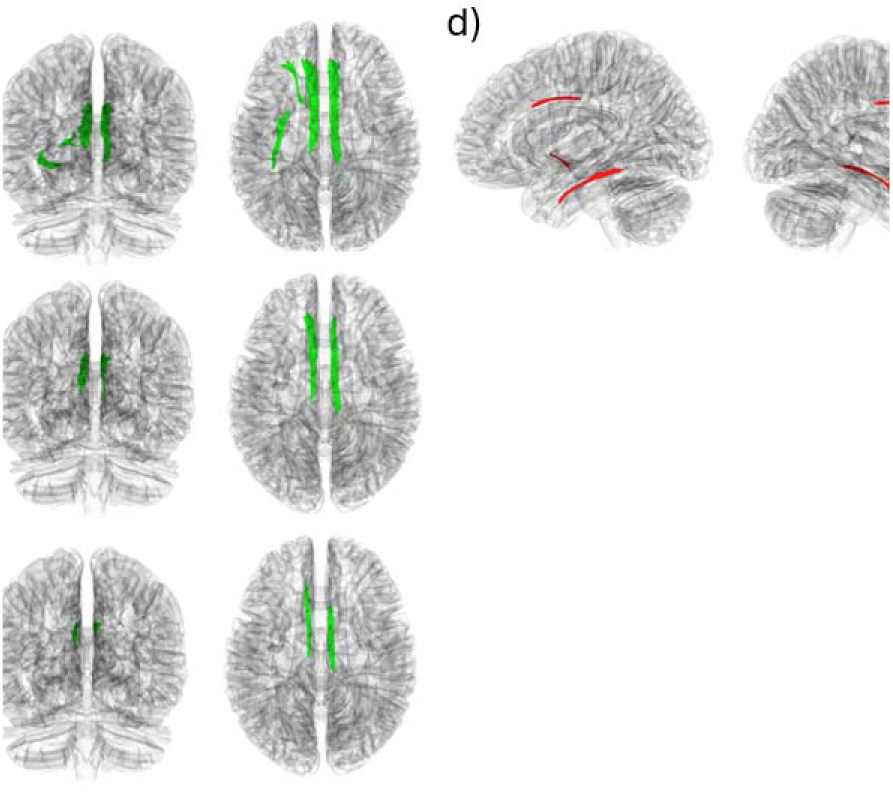
Restricted connectometry results of PACAP associations with white matter fractional anisotropy (FA), within participants scanned with the Adult Lifespan Protocol. Panels (a-c) show white matter segments (green) where higher PACAP levels correlated with higher FA at different T-thresholds: (a) T = 2.0; (b) T = 2.5; (c) T = 3.0. (d) Shows higher white matter segments (red) where higher PACAP levels correlated with lower white matter FA at T = 2.0, none remaining significant at higher T-thresholds.

**Figure S10.**
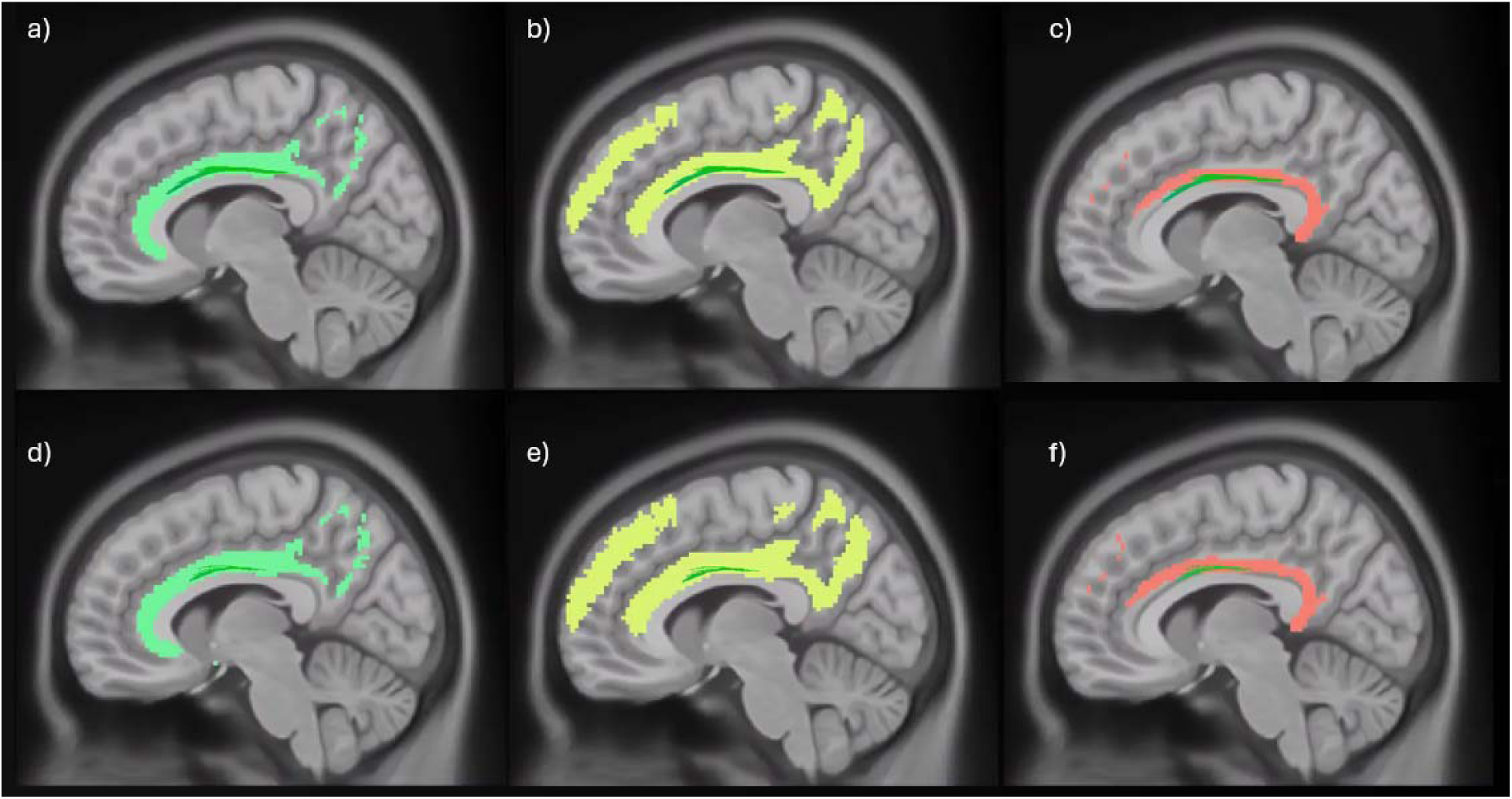
Overlap of white matter connectometry findings at *T* = 3.0 for cortisol and PACAP with major cingulum tract regions. Panels (a–c) show cortisol-associated streamlines (green) overlaid on reference masks for (a) the parolfactory cingulum, (b) the frontoparietal cingulum (yellow), and (c) the frontoparahippocampal cingulum (salmon). Panels (d–f) show PACAP-associated streamlines (green) overlaid on the same respective cingulum subregions: (d) parolfactory, (e) frontoparietal (yellow), and (f) frontoparahippocampal (salmon).

**Table S1.** Traumatic events reported by participants (N=131)

| **LEC-5 Event Type** | **Experienced N (%)** | **Witnessed N (%)** |
| --- | --- | --- |
| Natural disaster (for example, flood, hurricane, tornado, earthquake) | 44 (33.6%) | 24 (18.2%) |
| Fire or explosion | 15 (11.5%) | 33 (25.2%) |
| Transportation accident (for example, car accident, boat accident, train wreck, plane crash) | 75 (57.3%) | 40 (30.5%) |
| Serious accident at work, home, or during recreational activity | 25 (19.1%) | 37 (28.2%) |
| Exposure to toxic substance (for example, dangerous chemicals, radiation) | 10 (7.6%) | 6 (4.6%) |
| Physical assault (for example, being attacked, hit, slapped, kicked, beaten up) | 98 (74.8%) | 50 (38.2%) |
| Assault with a weapon (for example, being shot, stabbed, threatened with a knife, gun, bomb) | 39 (29.8%) | 33 (25.2%) |
| Sexual assault (rape, attempted rape, made to perform any type of sexual act through force or threat of harm) | 87 (66.4%) | 17 (13.0%) |
| Other unwanted or uncomfortable sexual experience | 106 (80.9%) | 20 (15.3%) |
| Combat or exposure to a warzone (in the military or as a civilian) | 5 (3.8%) | 6 (4.6%) |
| Captivity (for example, being kidnapped, abducted, held hostage, prisoner of war) | 8 (6.1%) | 4 (3.1%) |
| Life-threatening illness or injury | 18 (13.7%) | 52 (39.7%) |
| Severe human suffering | 19 (14.5%) | 35 (26.7%) |
| Sudden violent death (for example, homicide, suicide) | 8 (6.1%) | 35 (26.7%) |
| Sudden accidental death | 11 (8.4%) | 29 (22.1%) |
| Serious injury, harm, or death you caused to someone else | 12 (9.2%) | 9 (6.9%) |
| Any other very stressful event or experience | 51 (38.9%) | 16 (12.2%) |

*Note:* LEC-5: Life Events Checklist for DSM-5. Eight participants did not complete the LEC-5.

**Table S2.**
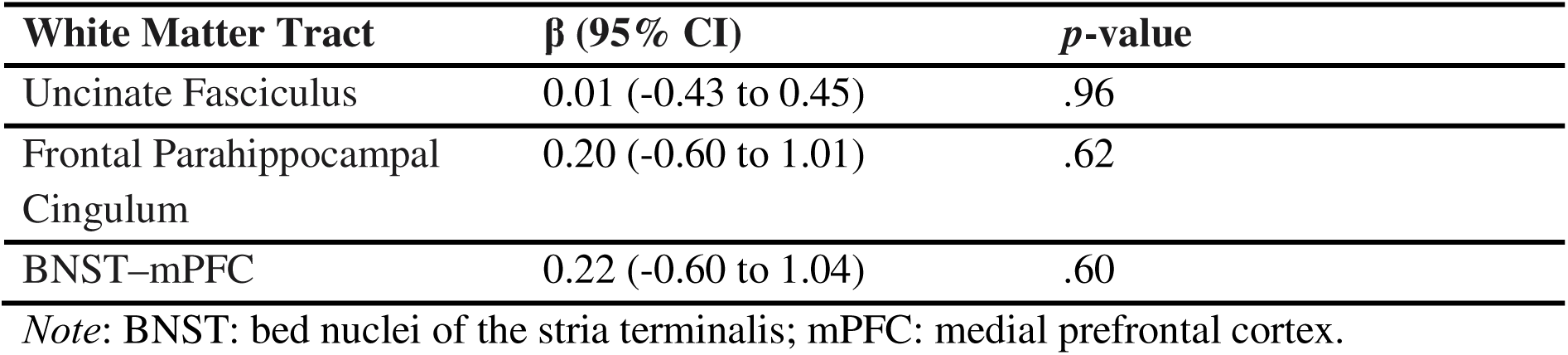
Cortisol-by-hemisphere interaction terms from linear mixed-effects models evaluating associations between cortisol and tract-average fractional anisotropy in three white matter tracts of interest.

**Table S3.** PACAP-by-hemisphere interaction terms from linear mixed-effects models evaluating associations between PACAP and tract-average fractional anisotropy in three white matter tracts of interest.

| White Matter Tract | $\beta$ (95% CI) | *p*-value |
| --- | --- | --- |
| Uncinate Fasciculus | 0.0015 (-0.002 to 0.004) | .33 |
| Frontal Parahippocampal<br>Cingulum | -0.0004 (-0.006 to 0.005) | .90 |
| BNST–mPFC | -0.0018 (-0.007 to -0.004) | .53 |

*Note:* BNST: bed nuclei of the stria terminalis; mPFC: medial prefrontal cortex.

## Notes

### Competing Interest Statement

The authors have declared no competing interest.

### Author Declarations

The Mass General Brigham Human Research Committee gave ethical approval for this work.

### Summary of Updates

This revision corrects an error in a grant number within the Funding disclosure section.

